# Unmasking Disparities in Caesarean Section Utilization in Nigeria: A K-Means Cluster Analysis of Obstetric Risk Profiles Using Machine Learning Approach

**DOI:** 10.64898/2026.04.30.26352131

**Authors:** Samuel Ayoola Ajeboriogbon, Abolaji Moses Ogunetimoju, Oluwafemi Lawal Bisiriyu

**Affiliations:** Department of Statistics, Obafemi Awolowo University, Ile-Ife, Osun State, Nigeria

**Keywords:** Caesarean section, K-means clustering, machine learning, health disparities, Nigeria, obstetric care, maternal health, Demographic and Health Survey

## Abstract

Caesarean section rates in Nigeria remain suboptimal, with significant disparities across socioeconomic and geographic strata. The objective of this research is to identify and characterize distinct obstetric risk profiles associated with caesarean section utilization in Nigeria using K-means cluster analysis, and to examine the sociodemographic and geographic factors driving these disparities. We analyzed data from 13,915 women with recent births in the 2024 Nigeria Demographic and Health Survey. Fourteen variables spanning demographics, socioeconomic status, healthcare access, medical history, and geography were used as clustering features. K-Means clustering was performed with optimal cluster selection based on silhouette score, Davies Bouldin index, and Calinski Harabasz index. Bootstrap validation with 100 iterations assessed cluster stability, while chi-square tests and logistic regression examined associations between cluster membership and surgical delivery. Ten distinct clusters were identified with rates ranging from 1.7% to 14.4%, representing an 8.4-fold variation. The highest utilization cluster at 14.4% comprised urban, highly educated, wealthy women with extensive antenatal care averaging 16.5 visits, while the lowest utilization cluster at 1.7% consisted of rural, poorly educated, impoverished women with minimal healthcare access averaging 2.3 visits. Cluster membership was significantly associated with utilization, and bootstrap analysis confirmed cluster stability with a mean silhouette of 0.220. Machine learning based clustering reveals profound disparities in utilization across distinct population subgroups in Nigeria, highlighting the dual challenge of underutilization among disadvantaged rural populations and potential overutilization among urban elites. Targeted interventions addressing geographic, economic, and healthcare access barriers are essential to optimize utilization across all population segments.

## 1.0 INTRODUCTION

Caesarean section (CS) is a critical obstetric intervention that prevents maternal and neonatal mortality and morbidity when appropriately indicated (Betrán et al., 2018). The World Health Organization recommends population-level CS rates between 10% and 15% for optimal outcomes, with lower rates suggesting unmet needs for emergency obstetric care (WHO, 2015). However, profound global inequities exist; rates range from under 5% in many sub-Saharan African countries to over 50% in some high-income settings (Boerma et al., 2018). Nigeria, with over 200 million inhabitants, exemplifies this paradox. Despite contributing approximately 23% of global maternal deaths, its national surgical delivery rate remains critically low (WHO, 2019). The 2018 Nigeria Demographic and Health Survey recorded a 2.7% rate, reflecting substantial underutilization (National Population Commission & ICF, 2019). This aggregate obscure significant heterogeneity across geopolitical zones, urban and rural residences, wealth quintiles, and educational attainment (Adewuyi et al., 2019). Furthermore, Nigeria’s tiered healthcare system concentrates surgical services in secondary and tertiary facilities (Federal Ministry of Health, 2016). The maldistribution of these facilities, compounded by financial barriers, transportation, and sociocultural factors, creates substantial inequities in access to care (Okonofua et al., 2018).

Extensive literature identifies consistent predictors of CS utilization in Nigeria, including maternal age, parity, wealth, education, urban residence, and antenatal care (Adewuyi et al., 2019; Cavallaro et al., 2013; Osonwa et al., 2016). A strong positive association exists between socioeconomic advantage and utilization, with women in the richest wealth quintiles showing several-fold higher rates than the poorest (Boerma et al., 2018). Healthcare factors like facility-based delivery, skilled birth attendants, and health insurance also increase utilization (Ronsmans et al., 2006). Geographic analyses reveal stark disparities, with southern Nigerian regions demonstrating higher rates due to better infrastructure and differing sociocultural norms (Adewuyi et al., 2019). While clinical indications like hypertensive disorders and diabetes influence decisions (Betrán et al., 2018), the contribution of medical versus non-medical factors remains contested in resource-limited settings (Manyeh et al., 2018). Concerns persist that socioeconomic advantage may override clinical considerations (Manyeh et al., 2018).

Consequently, Nigeria faces a dual challenge: underutilization among disadvantaged women drives preventable deaths, while potential overutilization among privileged subgroups raises concerns about unnecessary surgeries (Betran et al., 2016). To contextualize this, this study utilizes the Three Delays Model originally proposed by Thaddeus and Maine (1994), which identifies delays in deciding to seek care, reaching facilities, and receiving adequate care. Extended to explain CS utilization, this framework highlights how disadvantaged women face compounded delays resulting in underutilization even when medically necessary. Conversely, women with high socioeconomic status and urban residence may face minimal delays, facilitating surgical access without strict medical indications. This approach is further supported by the social determinants of health framework (Solar & Irwin, 2010), which emphasizes how structural factors shape intermediary healthcare behaviors and access.

Despite existing research, traditional regression approaches often evaluate predictors independently, inadequately capturing the multidimensional nature of obstetric risk profiles and missing critical intersections. Cluster analysis addresses this gap by identifying homogeneous subgroups sharing similar multidimensional profiles (Everitt et al., 2011). While clustering has identified risk phenotypes for adverse outcomes (Sovio et al., 2020), its application to CS utilization remains limited. Thus, the primary objective of this study is to identify and characterize distinct population subgroups with homogeneous obstetric risk profiles regarding CS utilization in Nigeria using K-means cluster analysis. Specific objectives include determining the optimal number of clusters based on sociodemographic, medical, and geographic traits; characterizing utilization rates; examining associations with delivery methods while adjusting for confounders; and interpreting findings for public health policy.

This research contributes significantly by applying machine learning clustering techniques to maternal health in Nigeria, demonstrating its utility in identifying distinct risk profiles. By analyzing the full spectrum of utilization within a unified framework, this study provides actionable insights into Nigeria’s dual policy challenges. Identifying distinct clusters shifts focus from one-size-fits-all policies to targeted interventions. Clusters characterized by underutilization can be prioritized for access improvements, while those with potential overutilization can benefit from evidence-based surgical guidelines. Utilizing recent, nationally representative data from the 2024 demographic survey, this study offers contemporary estimates for policy planning and a methodological template for analyzing global healthcare utilization disparities.

## 2.0 METHODOLOGY

This study utilized a cross-sectional analytical design, analyzing secondary data from the nationally representative 2024 Nigeria Demographic and Health Survey (NDHS). The research applied unsupervised machine learning, specifically K-means clustering, alongside traditional inferential statistics to categorize distinct obstetric risk profiles across five interconnected analytical stages. The study setting encompasses Nigeria’s 36 states and the Federal Capital Territory, a highly populated nation marked by significant regional, maternal health, and socioeconomic disparities. In this system, Caesarean sections are largely restricted to secondary and tertiary healthcare facilities, which creates substantial geographic access barriers for women.

The 2024 NDHS utilized a stratified two-stage cluster sampling method to collect initial data from 39,050 women aged 15 to 49 through standardized, rigorously monitored face-to-face interviews. For the analysis, fourteen clustering features were specifically chosen based on the Three Delays Model and the social determinants of health framework. The primary outcome variable was a binary indicator denoting whether a Caesarean section delivery occurred for the most recent birth within a five-year recall period.

Rigorous data cleaning addressed substantial missing data, which primarily resulted from the survey structure restricting certain questions to women with recent births. To handle this, researchers created missingness indicators and imputed median and modal values for continuous and categorical variables, respectively. Cases missing the primary outcome or remaining feature data were excluded entirely, reducing the final analytic sample to 13,915 women. Finally, to guarantee equal feature contribution to distance calculations during clustering, all variables underwent Min-Max normalization to a standardized scale prior to analysis.

### K-Means Cluster Analysis

K-means clustering is an unsupervised machine learning algorithm that partitions observations into *k* distinct, non-overlapping clusters based on feature similarity (MacQueen, 1967). The algorithm minimizes the within-cluster sum of squares (WCSS), also known as inertia:

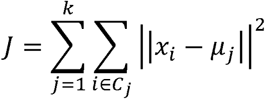

Where:

*J* = total within-cluster variance (inertia)

*k* = number of clusters

*C_j_* = set of observations in cluster *j x_i_*= observation *i*

μ*_j_* = centroid of cluster *j*

||·||² = squared Euclidean distance

## Optimal “k” Selection

The optimal number of clusters was determined using multiple validation metrics:

1. Silhouette Score:

The silhouette coefficient measures how similar observations are to their own cluster compared to other clusters (Rousseeuw, 1987):

Where:

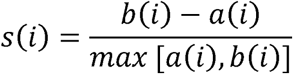

*a(i)* = average distance from observation *i* to other points in the same cluster

*b(i)* = minimum average distance from observation *i* to points in other clusters

The overall silhouette score is the mean across all observations, ranging from −1 to +1, with higher values indicating better-defined clusters.

2. Davies-Bouldin Index:

This index measures the ratio of within-cluster distances to between-cluster distances (Davies & Bouldin, 1979):

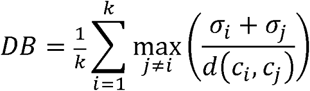

Where:

σ*_i_* = average distance of points in cluster *i* to centroid *c_i_ d(c_i_, c_j_)* = distance between centroids *i* and *j*

Lower values indicate better cluster separation.

3. Calinski-Harabasz Index:

This index measures the ratio of between-cluster dispersion to within-cluster dispersion (Caliński & Harabasz, 1974):

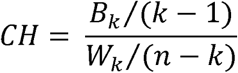

Where:

*B_k_* = between-cluster dispersion *W_k_*= within-cluster dispersion *k* = number of clusters

*n* = total observations

Higher values indicate better-defined clusters.

4. Elbow Method:

Within-cluster sum of squares (inertia) was plotted against k (2–14), with the “elbow point” indicating diminishing returns from additional clusters.

Bootstrap Stability Analysis

Cluster stability was assessed using bootstrap resampling (Efron & Tibshirani, 1993). One hundred bootstrap samples were drawn with replacement from the original dataset:

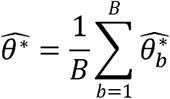

Where:

*B* = 100 bootstrap iterations

θ * = silhouette score from bootstrap sample *b*

The 95% confidence interval was computed using the percentile method:

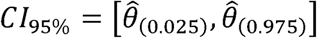

Narrow confidence intervals indicate stable cluster solutions.

### Caesarean Section Rate Calculation

CS rates were calculated for each cluster with 95% Wilson confidence intervals:

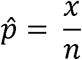

Where:

*x* = number of CS deliveries in cluster

*n* = total women in cluster

### Chi-Square Test for Association

The overall association between cluster membership and CS delivery was tested using Pearson’s chi-square test:

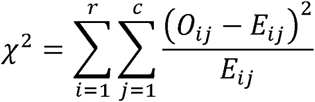

Where:

*O_ij_* = observed frequency in cell (*i*, *j*)

*E_ij_* = expected frequency under null hypothesis of independence

*r* = number of rows (clusters)

*c* = number of columns (CS status)

Effect size was quantified using Cramér’s V:

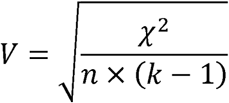

Where *k* = min (rows, columns). Values of 0.10, 0.30, and 0.50 represent small, medium, and large effects, respectively.

### Logistic Regression

Binary logistic regression models examined the association between cluster membership and CS delivery. The logistic model is specified as:

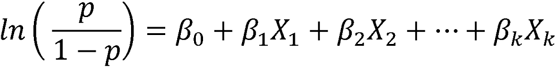

Where:

*p* = probability of CS delivery

β*_0_* = intercept

β*_1_…*β*_k_* = regression coefficients for cluster dummy variables and covariates Odds ratios were computed as:

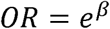

With 95% confidence intervals:

Two models were estimated:

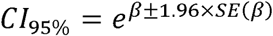

**1. Unadjusted model:** Cluster membership as sole predictor
**2. Adjusted model:** Cluster membership adjusted for age, wealth, education, and urban/rural residence

### Dimensionality Reduction (PCA)

Principal Component Analysis (PCA) was performed for visualization. PCA identifies orthogonal components that maximize variance:

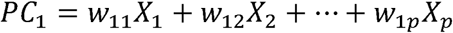

Where *w* represents the eigenvector weights (loadings) for each original variable.

### Ethical Considerations

This study utilized publicly available, de-identified secondary data from the DHS Program. The original NDHS 2024 survey received ethical approval from the National Health Research Ethics Committee of Nigeria and the ICF Institutional Review Board. Informed consent was obtained from all survey participants prior to data collection.

As a secondary analysis of anonymized data, this study did not require additional ethical approval. No individual participants can be identified from the dataset, and all analyses were conducted at the aggregate level.

## 3.0 RESULTS

Descriptive Analysis

Study Population Characteristics

The final analytic sample comprised 13,915 women with recent births and complete data on clustering features. Table 1 presents the baseline characteristics of the study population.

**Table 1:**
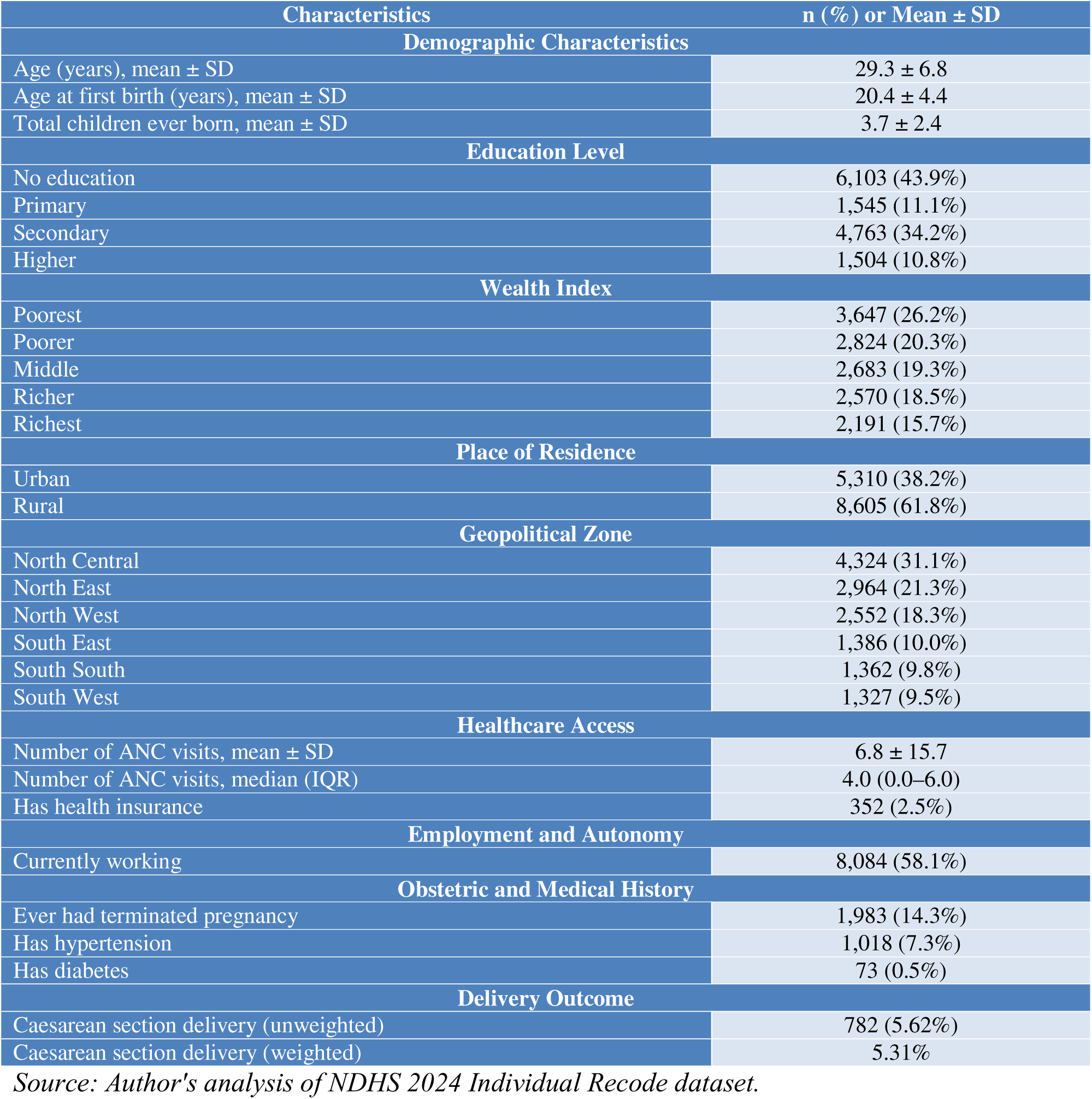
Baseline Characteristics of Study Population (N = 13,915)

| Characteristics | n (%) or Mean ± SD |
| --- | --- |
| **Demographic Characteristics** |  |
| Age (years), mean ± SD | 29.3 ± 6.8 |
| Age at first birth (years), mean ± SD | 20.4 ± 4.4 |
| Total children ever born, mean ± SD | 3.7 ± 2.4 |
| **Education Level** |  |
| No education | 6,103 (43.9%) |
| Primary | 1,545 (11.1%) |
| Secondary | 4,763 (34.2%) |
| Higher | 1,504 (10.8%) |
| **Wealth Index** |  |
| Poorest | 3,647 (26.2%) |
| Poorer | 2,824 (20.3%) |
| Middle | 2,683 (19.3%) |
| Richer | 2,570 (18.5%) |
| Richest | 2,191 (15.7%) |
| **Place of Residence** |  |
| Urban | 5,310 (38.2%) |
| Rural | 8,605 (61.8%) |
| **Geopolitical Zone** |  |
| North Central | 4,324 (31.1%) |
| North East | 2,964 (21.3%) |
| North West | 2,552 (18.3%) |
| South East | 1,386 (10.0%) |
| South South | 1,362 (9.8%) |
| South West | 1,327 (9.5%) |
| **Healthcare Access** |  |
| Number of ANC visits, mean ± SD | 6.8 ± 15.7 |
| Number of ANC visits, median (IQR) | 4.0 (0.0–6.0) |
| Has health insurance | 352 (2.5%) |
| **Employment and Autonomy** |  |
| Currently working | 8,084 (58.1%) |
| **Obstetric and Medical History** |  |
| Ever had terminated pregnancy | 1,983 (14.3%) |
| Has hypertension | 1,018 (7.3%) |
| Has diabetes | 73 (0.5%) |
| **Delivery Outcome** |  |
| Caesarean section delivery (unweighted) | 782 (5.62%) |
| Caesarean section delivery (weighted) | 5.31% |

*Source: Author's analysis of NDHS 2024 Individual Recode dataset.*

The study population had a mean age of 29.3 years (SD = 6.8), with mean age at first birth of 20.4 years (SD = 4.4) and mean parity of 3.7 children (SD = 2.4). Nearly half of participants (43.9%) had no formal education, while 45.0% had secondary or higher education. The wealth distribution was skewed toward lower quintiles, with 46.5% in the poorest or poorer categories. The majority (61.8%) resided in rural areas, and the northern geopolitical zones (North Central, North East, North West) accounted for 70.7% of the sample.

Healthcare access indicators revealed substantial gaps: mean ANC visits was 6.8 but with high variability (SD = 15.7), and only 2.5% had health insurance coverage. Medical conditions were relatively uncommon, with 7.3% reporting hypertension and 0.5% reporting diabetes.

The overall CS rate was 5.62% (unweighted) and 5.31% (weighted), corresponding to 782 CS deliveries among 13,915 births.

Core Findings

### Cluster Validation and Selection

K-means clustering was performed for k = 2 through k = 14 clusters. Table 2 presents validation metrics across cluster solutions.

**Table 2:** Cluster Validation Metrics Across k Values.

| k | Inertia | Silhouette Score | Davies-Bouldin Index | Calinski-Harabasz Index |
| --- | --- | --- | --- | --- |
| 2 | 22,992 | 0.226 | 1.840 | 3,911 |
| 3 | 20,663 | 0.196 | 1.956 | 2,960 |
| 4 | 18,908 | 0.175 | 2.041 | 2,587 |
| 5 | 17,494 | 0.177 | 1.930 | 2,378 |
| 6 | 16,717 | 0.174 | 1.835 | 2,120 |
| 7 | 15,677 | 0.191 | 1.781 | 2,037 |
| 8 | 14,943 | 0.197 | 1.689 | 1,929 |
| 9 | 13,921 | 0.219 | 1.593 | 1,940 |
| 10 | 13,453 | 0.218 | 1.556 | 1,838 |
| 11 | 13,090 | 0.220 | 1.615 | 1,738 |
| 12 | 12,529 | 0.218 | 1.553 | 1,707 |
| 13 | 12,207 | 0.235 | 1.591 | 1,637 |
| 14 | 11,930 | 0.238 | 1.572 | 1,571 |

*Source: Author's analysis of NDHS 2024 data.*

**Table 3:** Comparison of k = 2 and k = 10 Solutions.

| Metric | k = 2 | k = 10 |
| --- | --- | --- |
| Silhouette Score | 0.226 | 0.227 |
| Davies-Bouldin Index | 1.840 | 1.666 |
| Calinski-Harabasz Index | 3,911 | 1,838 |
| Cramér's V (CS association) | 0.104 | 0.180 |
| CS rate range | 3.5%–8.4% | 1.7%–14.4% |

*Source: Author's computations from NDHS 2024 data.*

**Table 4:** Caesarean Section Rates by Cluster (k = 10) with 95% confidence intervals.

| Cluster | n (%) | CS Cases | CS Rate (%) | Weighted Rate (%) | 95% CI |
| --- | --- | --- | --- | --- | --- |
| 0 | 1,577 (11.3%) | 34 | 2.16 | 2.18 | 1.55–3.00 |
| 1 | 817 (5.9%) | 51 | 6.24 | 5.54 | 4.78–8.11 |
| 2 | 1,793 (12.9%) | 41 | 2.29 | 2.57 | 1.69–3.09 |
| 3 | 1,085 (7.8%) | 156 | 14.38 | 14.39 | 12.42–16.59 |
| 4 | 914 (6.6%) | 83 | 9.08 | 8.96 | 7.39–11.12 |
| 5 | 1,883 (13.5%) | 47 | 2.50 | 2.04 | 1.88–3.30 |
| 6 | 1,295 (9.3%) | 114 | 8.80 | 8.64 | 7.38–10.47 |
| 7 | 2,318 (16.7%) | 40 | 1.73 | 1.60 | 1.27–2.34 |
| 8 | 1,008 (7.2%) | 105 | 10.42 | 10.83 | 8.68–12.46 |
| 9 | 1,225 (8.8%) | 111 | 9.06 | 9.11 | 7.58–10.80 |
| **Total** | **13,915** | **782** | **5.62** | **5.31** | — |

*Source: Author's computations from NDHS 2024 data.*

**Figure 1:**
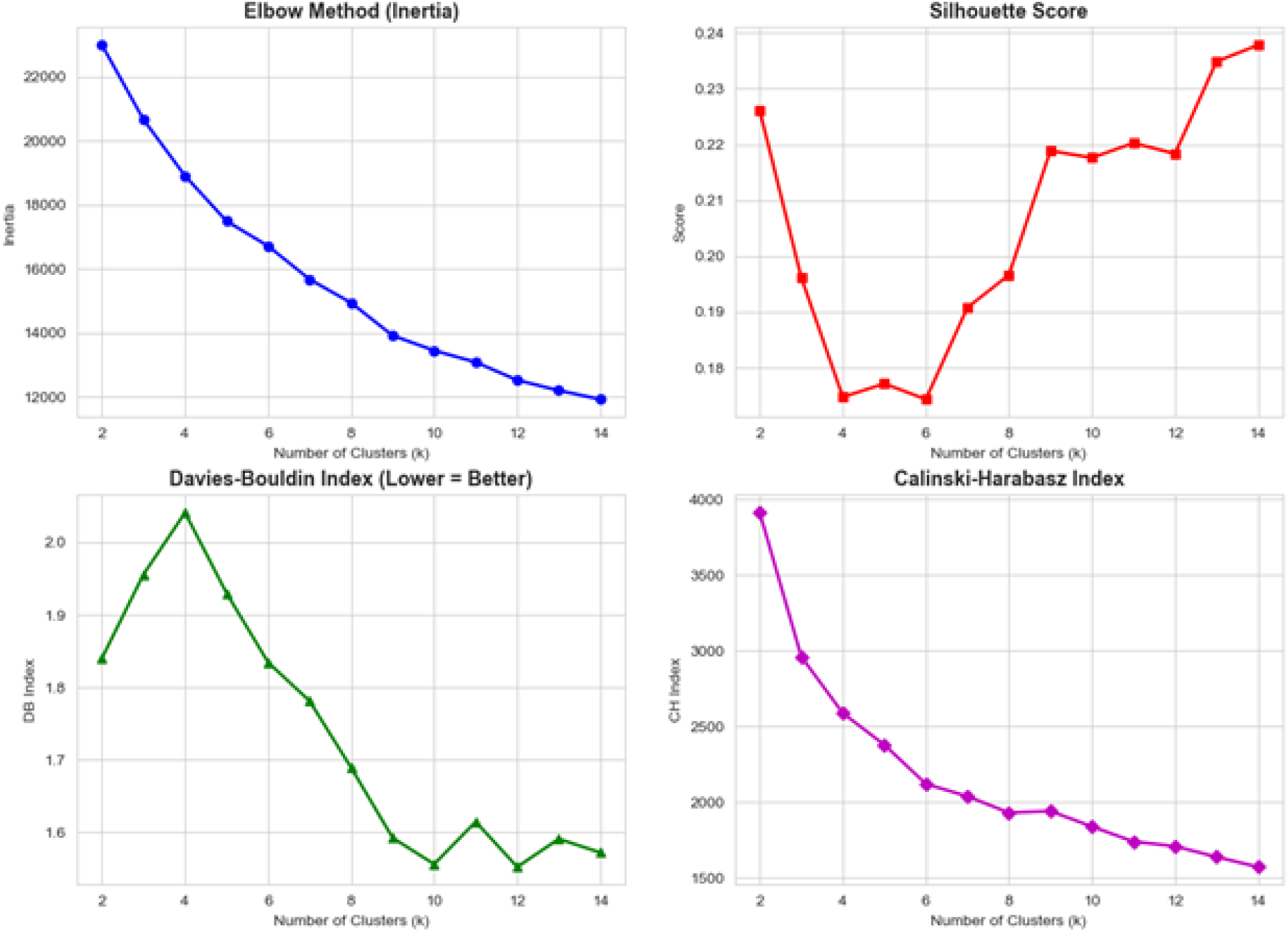
Cluster Validation Metrics Across k Values. Panel A shows the Elbow Method (Inertia), Panel B shows Silhouette Scores, Panel C shows Davies-Bouldin Index, and Panel D shows Calinski-Harabasz Index for k = 2 to 14 clusters. The elbow method suggested diminishing returns beyond k = 5–6 clusters. Silhouette scores were relatively stable across solutions, ranging from 0.175 to 0.238. The Davies-Bouldin index favored higher k values (lower is better), while the Calinski-Harabasz index favored k = 2 (higher is better), reflecting its preference for parsimonious solutions. We compared k = 2 and k = 10 solutions in detail (Table 3), as k = 2 demonstrated optimal statistical metrics while k = 10 offered greater clinical granularity.

The **k = 10 solution was selected** based on: (1) comparable silhouette score (0.227 vs. 0.226), (2) superior Davies-Bouldin index (1.67 vs. 1.84), (3) substantially stronger association with CS utilization (Cramér’s V = 0.18 vs. 0.10), and (4) capacity to reveal clinically meaningful gradations in obstetric risk profiles, with CS rates ranging 8.4-fold compared to 2.4-fold for k = 2.

### Bootstrap Stability Analysis

**Figure 2:**
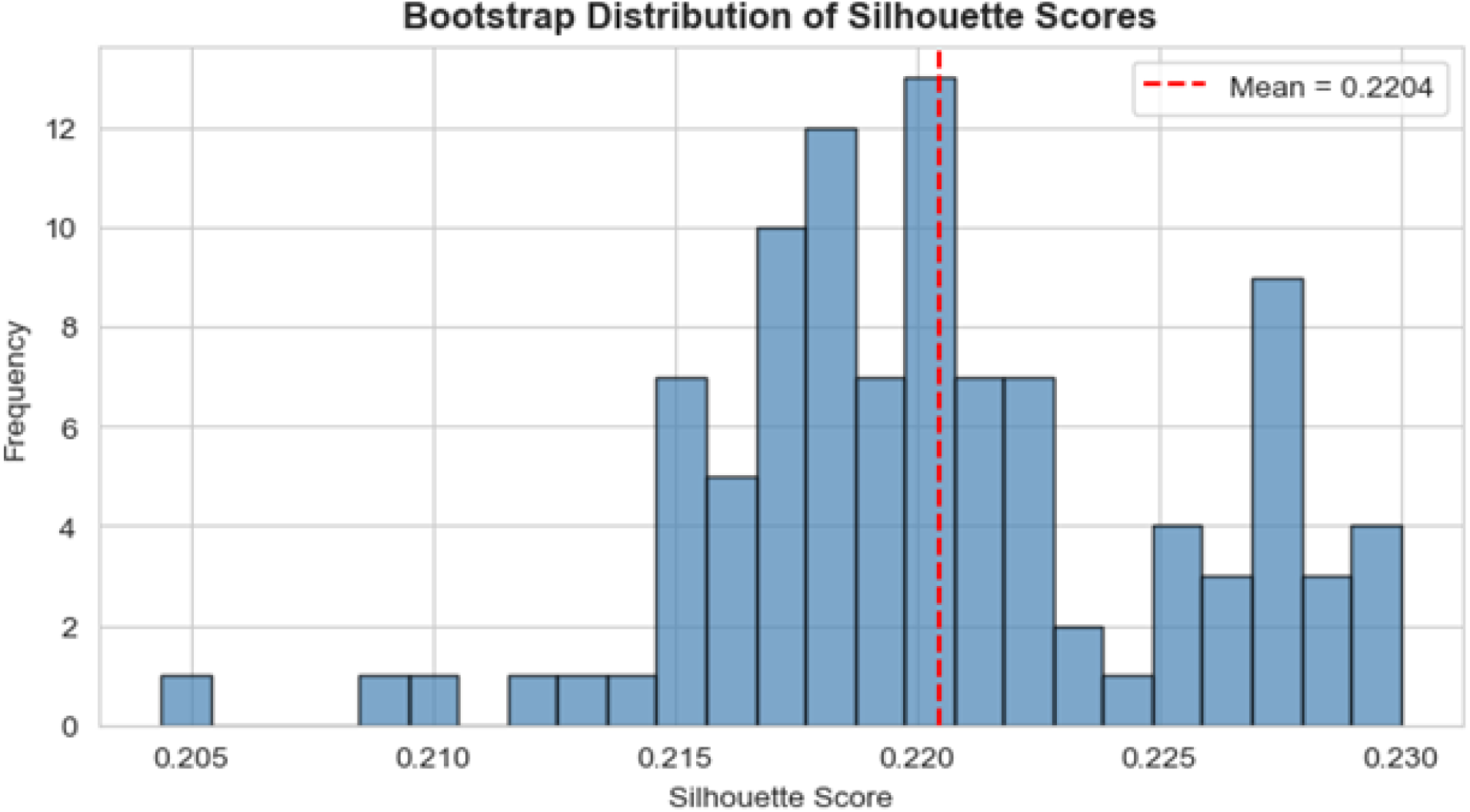
Bootstrap Distribution of Silhouette Scores (k = 10). Histogram showing the distribution of silhouette scores across 100 bootstrap iterations. The red dashed line indicates the mean silhouette score (0.220) with 95% confidence interval, 0.211–0.229. The narrow confidence interval indicates that cluster structure is robust and not unduly influenced by sampling variability.

**Figure 3:**
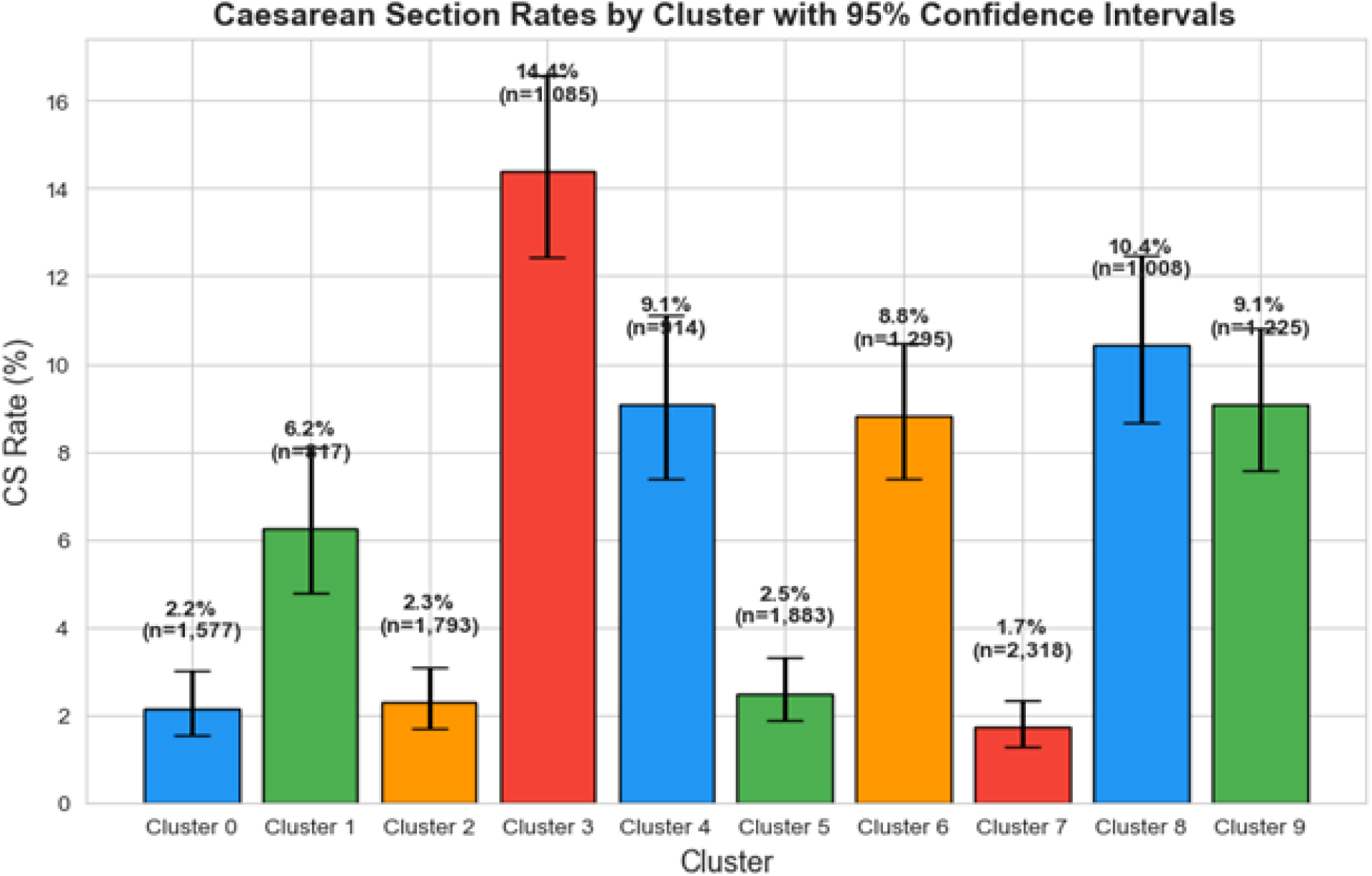
Caesarean Section Rates by Cluster with 95% Confidence Intervals. Bar chart showing CS rates across 10 clusters with error bars representing 95% confidence intervals. Rates ranged from 1.7% (Cluster 7) to 14.4% (Cluster 3), representing an 8.4-fold difference. CS rates varied substantially across clusters, ranging from 1.73% in Cluster 7 (lowest) to 14.38% in Cluster 3 (highest), representing an 8.4-fold difference. Four clusters (3, 4, 8, 9) demonstrated CS rates exceeding the national average, while four clusters (0, 2, 5, 7) had rates below 3%.

#### Cluster Characteristics and CS Rates

Table 5 presents detailed cluster profiles across sociodemographic, healthcare access, and medical characteristics.

**Table 5:**
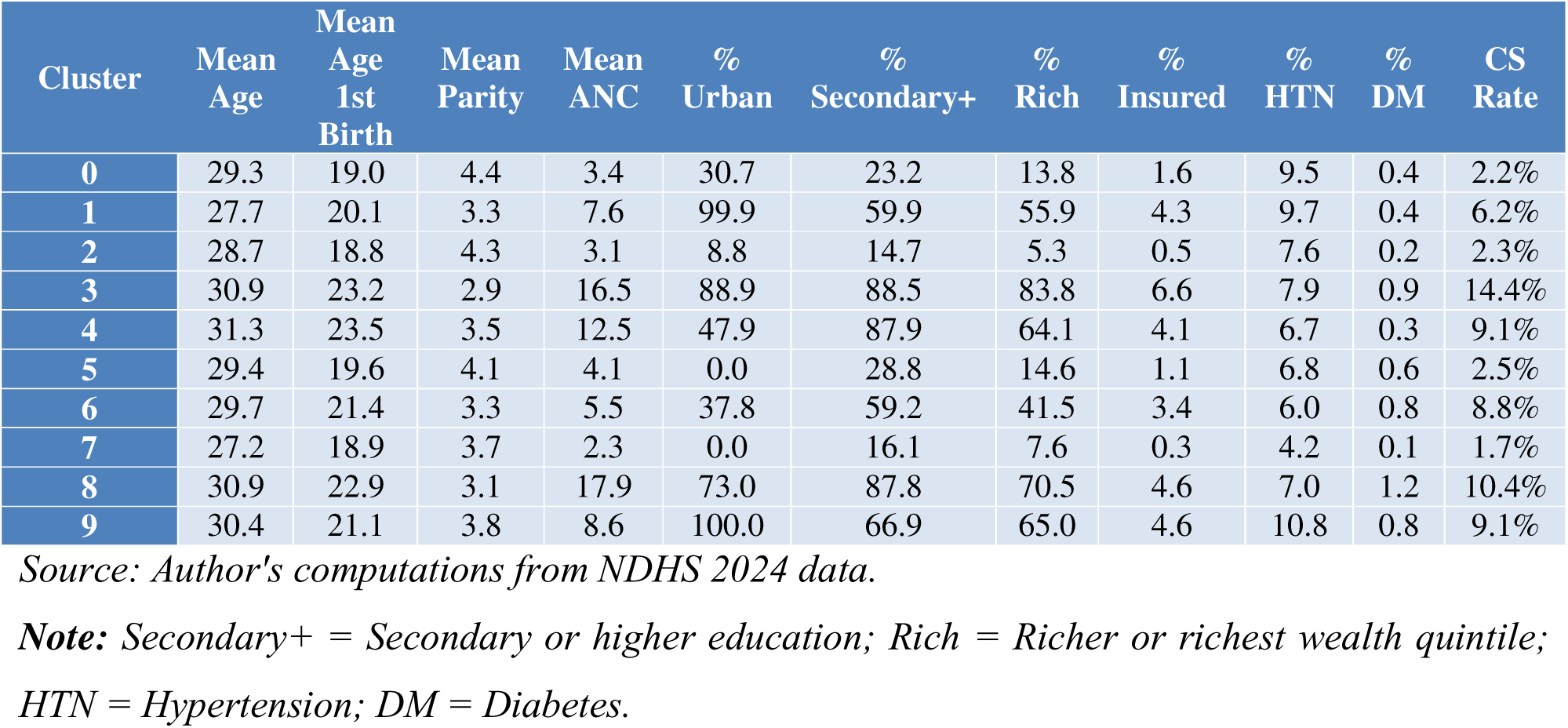
Cluster Profiles - Sociodemographic and Clinical Characteristics.

**Figure 4:** Cluster Profiles Radar Chart (Scaled Feature Means). Radar chart displaying scaled mean values for age, age at first birth, education, wealth, parity, and ANC visits across 10 clusters. High-CS clusters (3, 8, 4) demonstrate elevated education, wealth, and ANC utilization compared to low-CS clusters (7, 2, 0).

#### Cluster Interpretation

Based on the profiling results, Table 6 provides clinical interpretation and naming of clusters, ordered by CS rate.

**Table 6:**
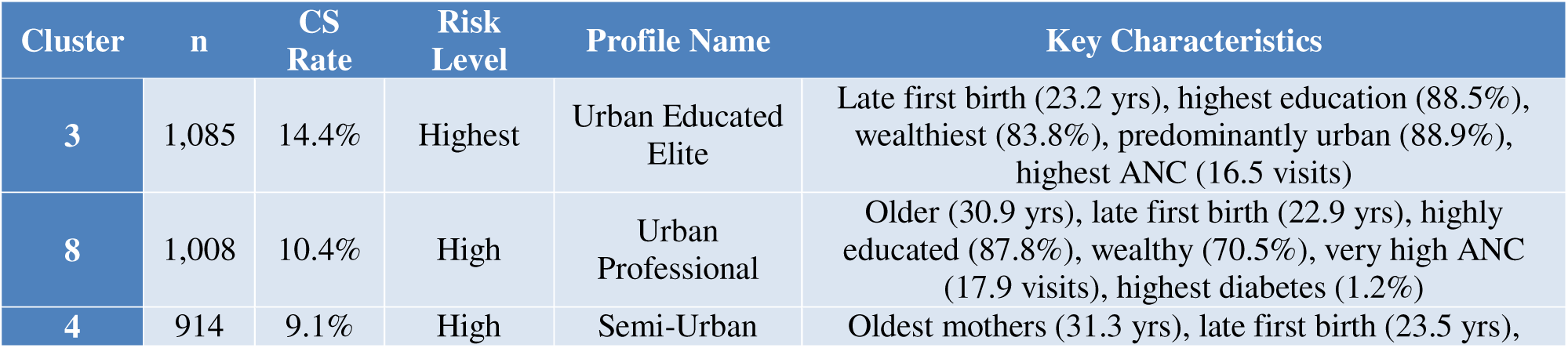
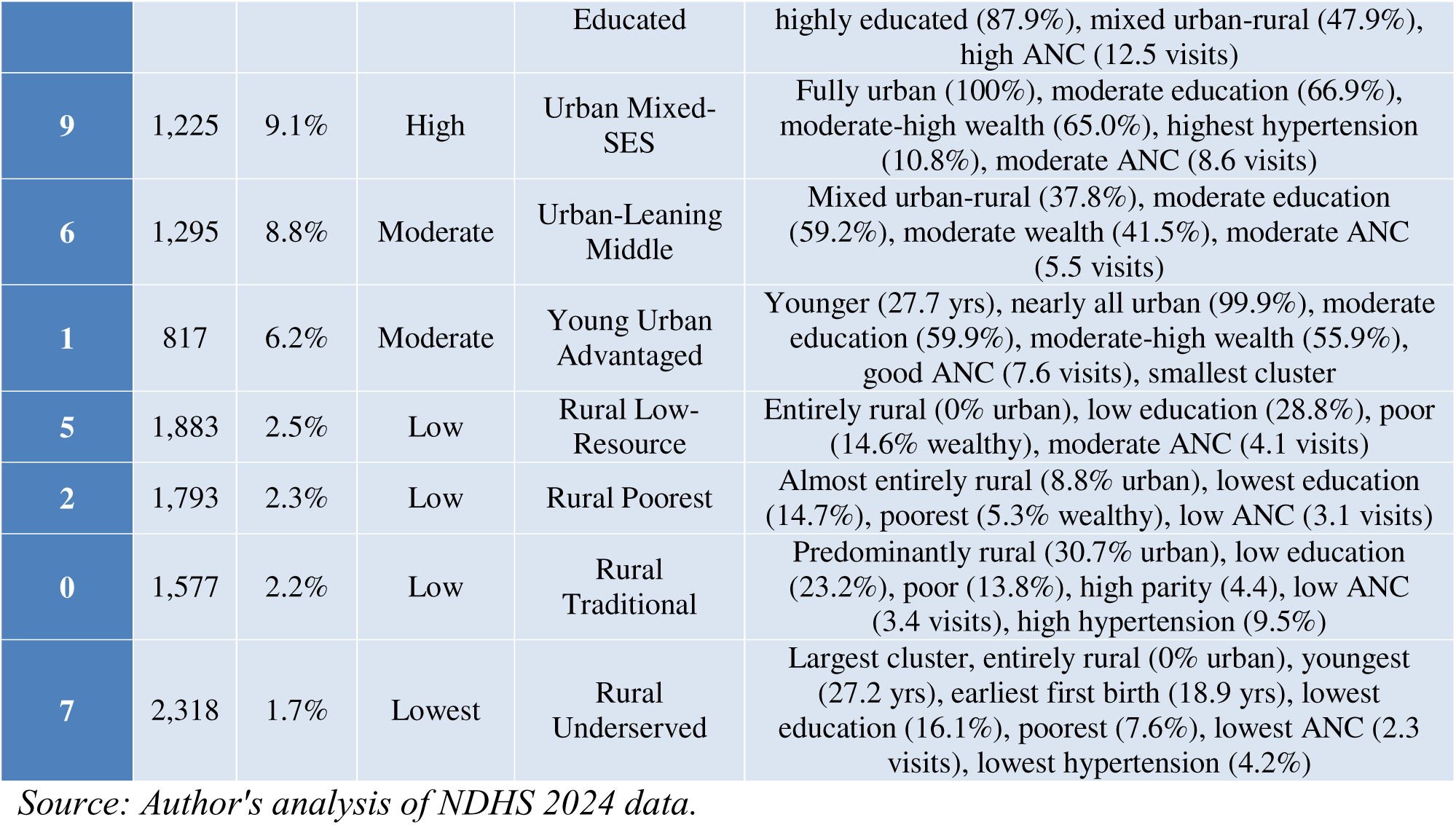
Clinical Cluster Interpretation and Risk Profiles.

The clusters revealed a clear **socioeconomic gradient** in CS utilization. High-CS clusters (3, 8, 4, 9) shared characteristics of urban or peri-urban residence, high educational attainment, wealth, and extensive antenatal care utilization. Low-CS clusters (7, 2, 0, 5) were characterized by rural residence, limited education, poverty, and minimal healthcare access.

Notably, **Cluster 7 (Rural Underserved)** represented the largest single cluster (16.7% of the sample) with the lowest CS rate (1.7%), while **Cluster 3 (Urban Educated Elite)** had the highest CS rate (14.4%), exceeding WHO recommendations for population-level CS rates.

#### Statistical Association Testing Chi-Square Test

The overall association between cluster membership and CS delivery was statistically significant:

- χ² = 448.12
- Degrees of freedom = 9
- p-value < 0.001
- Cramér’s V = 0.180

The Cramér’s V of 0.18 indicates a small-to-medium effect size, suggesting that cluster membership explains a meaningful proportion of variance in CS utilization.

#### Post-Hoc Pairwise Comparisons

Table 7 presents selected pairwise comparisons with Bonferroni-corrected p-values.

**Table 7:** Pairwise Post-Hoc Comparisons (Selected Significant Results)

| Comparison | $\chi^2$ |
| --- | --- |
| Cluster 3 vs 7 | 215.64 |
| Cluster 2 vs 3 | 153.09 |
| Cluster 3 vs 5 | 150.66 |
| Cluster 0 vs 3 | 143.02 |
| Cluster 7 vs 8 | 125.19 |
| Cluster 7 vs 9 | 103.91 |
| Cluster 4 vs 7 | 94.88 |
| Cluster 2 vs 8 | 84.68 |

Source: Author's computations from NDHS 2024 data.
Note: \* $p < 0.001$ after Bonferroni correction.

Of 45 pairwise comparisons, 31 (68.9%) were statistically significant after Bonferroni correction, confirming that most clusters differ meaningfully in CS utilization rates.

#### Logistic Regression

Table 8 presents unadjusted and adjusted logistic regression results with Cluster 7 (lowest CS rate) as the reference category.

**Table 8:** Logistic Regression - Association Between Cluster Membership and CS Delivery.

| Cluster | Unadjusted OR | 95% CI | p-value | Adjusted OR | 95% CI | p-value |
| --- | --- | --- | --- | --- | --- | --- |
| Cluster 7 (ref) | 1.00 | — | — | 1.00 | — | — |
| Cluster 0 | 1.25 | 0.79–1.99 | 0.335 | 0.88 | 0.54–1.42 | 0.600 |
| Cluster 1 | 3.79 | 2.49–5.78 | <0.001 | 0.94 | 0.57–1.53 | 0.795 |
| Cluster 2 | 1.33 | 0.86–2.07 | 0.201 | 1.50 | 0.96–2.35 | 0.078 |
| Cluster 3 | 9.56 | 6.70–13.65 | <0.001 | 1.27 | 0.82–1.96 | 0.285 |
| Cluster 4 | 5.69 | 3.87–8.36 | <0.001 | 0.97 | 0.63–1.49 | 0.880 |
| Cluster 5 | 1.46 | 0.95–2.23 | 0.083 | 0.90 | 0.58–1.40 | 0.646 |
| **Cluster 6** | 5.50 | 3.81–7.93 | <0.001 | 1.41 | 0.93–2.12 | 0.104 |
| **Cluster 8** | 6.62 | 4.56–9.61 | <0.001 | 1.00 | 0.64–1.55 | 0.988 |
| **Cluster 9** | 5.67 | 3.93–8.20 | <0.001 | 1.12 | 0.72–1.76 | 0.609 |

*Source: Author's computations from NDHS 2024 data.*
*Note: Adjusted for age (continuous), wealth index (continuous), education level (continuous), and urban/rural residence.*

**Table 9:** Cluster Distribution by Geopolitical Zone (%)

| Zone | Cluster 0 | Cluster 1 | Cluster 2 | Cluster 3 | Cluster 4 | Cluster 5 | Cluster 6 | Cluster 7 | Cluster 8 | Cluster 9 |
| --- | --- | --- | --- | --- | --- | --- | --- | --- | --- | --- |
| North Central | 16.6 | 7.6 | 0.0 | 3.2 | 0.1 | 23.5 | 0.0 | 40.7 | 0.0 | 8.3 |
| North East | 28.3 | 5.0 | 60.5 | 0.3 | 0.0 | 0.0 | 0.0 | 0.0 | 0.0 | 5.9 |
| North West | 0.0 | 6.3 | 0.0 | 0.0 | 0.0 | 20.0 | 50.7 | 14.8 | 0.0 | 8.1 |
| South East | 0.0 | 3.8 | 0.0 | 0.0 | 65.7 | 13.3 | 0.0 | 7.1 | 0.0 | 10.1 |
| South South | 0.1 | 5.6 | 0.0 | 0.0 | 0.0 | 6.2 | 0.0 | 3.3 | 74.0 | 10.7 |
| South West | 1.4 | 3.9 | 0.0 | **70.5** | 0.0 | 6.6 | 0.0 | 2.6 | 0.0 | 15.0 |

Source: Author's computations from NDHS 2024 data.
Note: **Bold values** indicate dominant cluster in each zone. Source: Author's computations from NDHS 2024 data.

In the unadjusted model, clusters with elevated CS rates demonstrated significantly increased odds of CS delivery compared to Cluster 7. Cluster 3 (Urban Educated Elite) showed the highest unadjusted odds ratio (OR = 9.56, 95% CI: 6.70–13.65), followed by Cluster 8 (OR = 6.62) and Cluster 4 (OR = 5.69).

However, after adjustment for age, wealth, education, and residence, all odds ratios attenuated substantially and became non-significant. The marked attenuation indicates that the cluster-CS associations are largely explained by the socioeconomic characteristics that differentiate the clusters, rather than representing independent cluster effects.

#### Principal Component Analysis

PCA revealed that the first two principal components explained 41.0% of the total variance:

- PC1: 27.5% variance explained
- PC2: 13.5% variance explained

Top loadings on PC1 included health decision autonomy (0.53), residence (0.39), education (0.32), wealth (0.32), and working status (0.30), suggesting this component primarily captures socioeconomic status and agency. PC2 loadings reflected age and parity dimensions.

**Figure 5:**
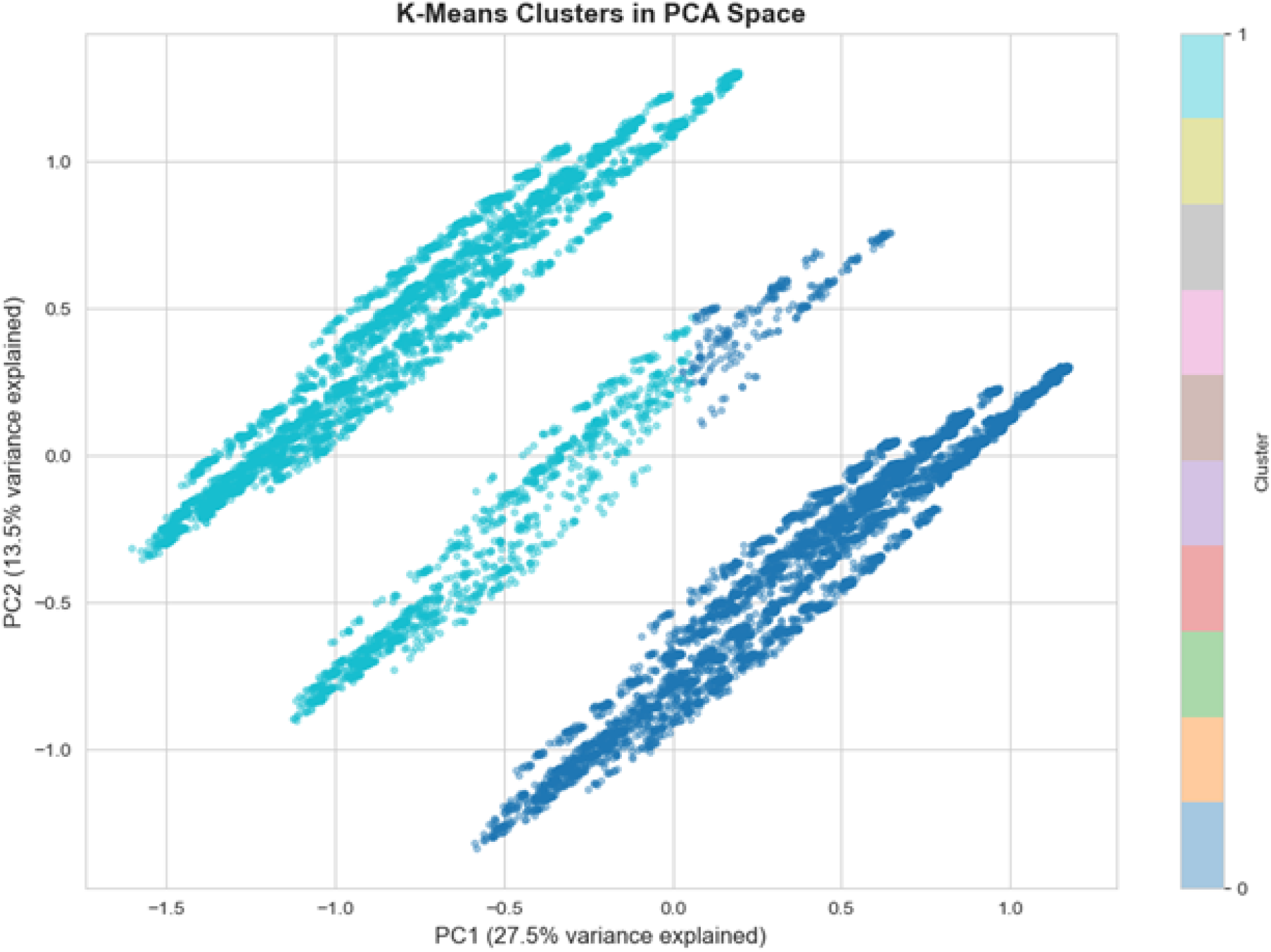
K-Means Clusters in PCA Space. Two-dimensional scatter plot showing the 10-cluster solution projected onto the first two principal components. PC1 (27.5% varianc explained) primarily reflects socioeconomic status and healthcare access, while PC2 (13.5% variance explained) captures age and parity dimensions. Points are colored by cluster assignment. Visualization of clusters in PCA space confirmed cluster separation, with high-CS clusters positioned at higher PC1 values (greater socioeconomic advantage) compared to low-CS clusters.

**Figure 6:**
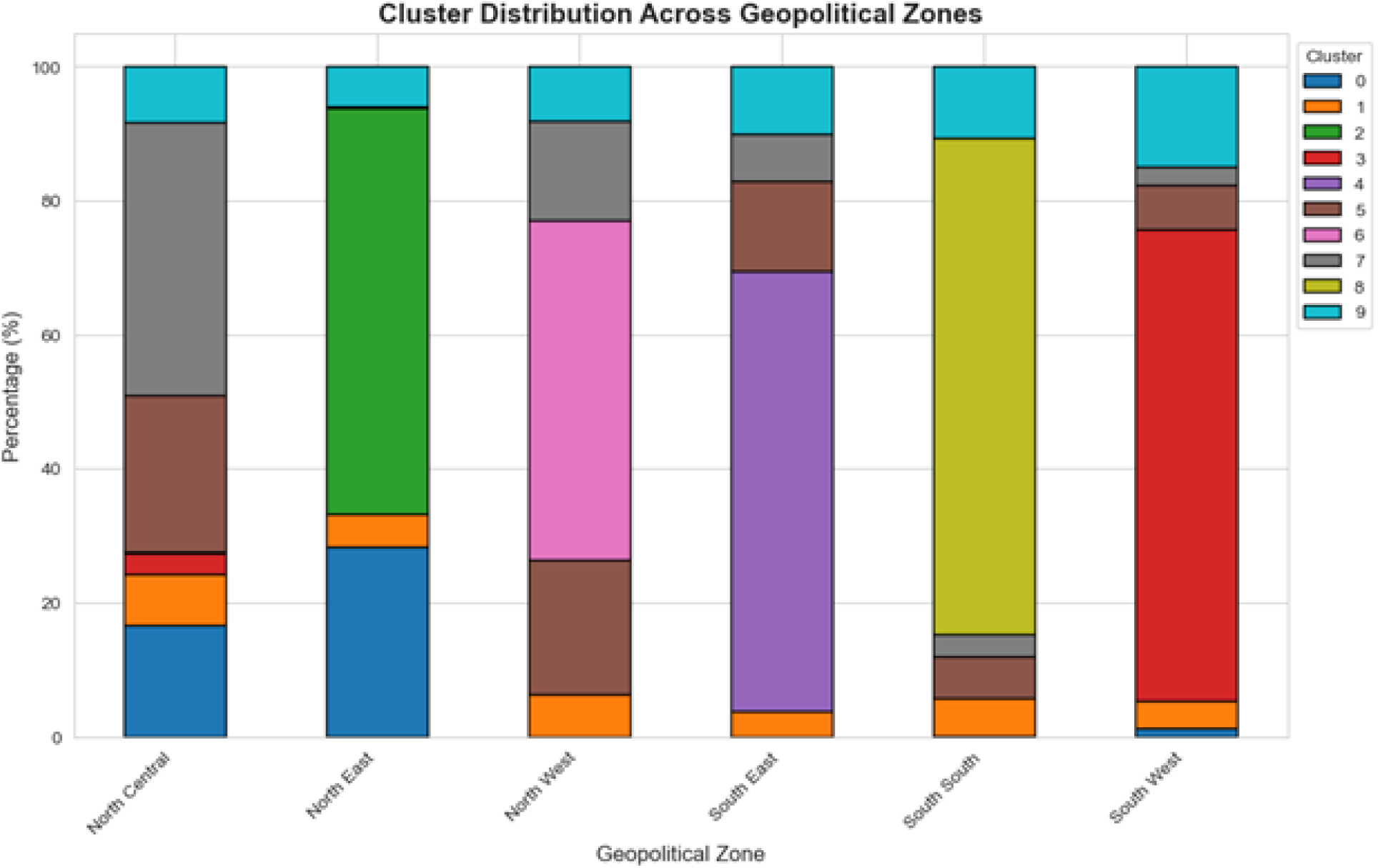
Cluster Distribution Across Geopolitical Zones. Stacked bar chart showing the percentage distribution of clusters across Nigeria’s six geopolitical zones. Each bar represents 100% of women within that zone, with colors indicating cluster membership (Clusters 0–9).

#### Geographic Distribution of Clusters

Analysis of cluster distribution across geopolitical zones revealed distinct geographic patterns that reflect the underlying socioeconomic and healthcare infrastructure disparities across Nigeria’s regions.

#### Key Findings

Each geopolitical zone is dominated by a single cluster, revealing a clear geographic segmentation of obstetric risk profiles:

| Zone | Dominant Cluster | % | Cluster CS Rate | Zone CS Profile |
| --- | --- | --- | --- | --- |
| North Central | Cluster 7 (Rural Underserved) | 40.7% | 1.7% | Low |
| North East | Cluster 2 (Rural Poorest) | 60.5% | 2.3% | **Lowest** |
| North West | Cluster 6 (Urban-Leaning Middle) | 50.7% | 8.8% | Moderate |
| South East | Cluster 4 (Semi-Urban Educated) | 65.7% | 9.1% | High |
| South South | Cluster 8 (Urban Professional) | 74.0% | 10.4% | High |
| South West | Cluster 3 (Urban Educated Elite) | 70.5% | 14.4% | **Highest** |

Source: Author's computations from NDHS 2024 data.

#### Interpretation

**Severe underutilization zones:** North Central (Cluster 7: 1.7% CS) and North East (Cluster 2: 2.3% CS) are dominated by the lowest-CS clusters, indicating critical unmet need for emergency obstetric care.

**Highest utilization zone:** South West is dominated by Cluster 3 (Urban Educated Elite) with 14.4% CS rate, approaching potential overutilization territory.

**Regional paradox in North West:** Despite being dominated by Cluster 6 (Urban-Leaning Middle, 8.8% CS), this zone’s actual CS rates may be lower due to healthcare infrastructure constraints not captured by sociodemographic clustering alone.

**Universal urban presence:** Cluster 9 (Urban Mixed-SES, 9.1% CS) appears in all zones (5.9%–15.0%), representing urban enclaves in state capitals across Nigeria.

**Southern advantage:** South East, South South, and South West are dominated by clusters with CS rates of 9.1%–14.4%, while North Central and North East are dominated by clusters with rates of 1.7%–2.3%, a 4-to-8-fold disparity driven by geography.

**Policy Implication:** Emergency obstetric care investments should prioritize North Central and North East zones, where over 60% of women belong to clusters with CS rates below 2.5%.

## 4.0 DISCUSSION

This study employed K-means cluster analysis to identify distinct population subgroups with homogeneous obstetric risk profiles associated with caesarean section utilization in Nigeria. The analysis revealed ten distinct clusters with CS rates ranging from 1.7% to 14.4%, representing an 8.4-fold variation that illuminates profound disparities obscured by aggregate national statistics.

The identified clusters demonstrate a clear socioeconomic gradient in CS utilization. High CS clusters comprising Clusters 3, 8, 4, and 9 represented 30.4% of the sample and shared characteristics of urban residence, high educational attainment, wealth, and extensive antenatal care utilization. Notably, Cluster 3 with a CS rate of 14.4% approaches the upper bound of WHO recommendations, raising questions about potential overutilization among this privileged subgroup. Conversely, low CS clusters comprising Clusters 7, 2, 0, and 5 represented 54.4% of the sample and were characterized by rural residence, limited education, poverty, and severely restricted healthcare access. These clusters demonstrated CS rates of 1.7% to 2.5%, far below the WHO recommended minimum of 10%.

The geographic analysis revealed stark regional segmentation. North East was dominated by Cluster 2 at 60.5% with a CS rate of 2.3%, while North Central was dominated by Cluster 7 at 40.7% with the lowest CS rate of 1.7%. In contrast, South West was dominated by Cluster 3 at 70.5% with the highest CS rate of 14.4%, and South South was dominated by Cluster 8 at 74.0% with a CS rate of 10.4%. This pattern exemplifies the dual challenge of “too little, too late” in Northern regions and potentially “too much, too soon” in Southern urban areas, consistent with findings documented across low and middle income countries by Miller and colleagues.

The attenuation of odds ratios after adjustment for socioeconomic factors provides important mechanistic insights. The finding that cluster CS associations were largely explained by age, wealth, education, and residence suggests that clusters represent distinct combinations of social determinants rather than independent risk phenotypes. This aligns with the Three Delays framework, where women in low CS clusters likely experience compounded delays in deciding to seek care, reaching facilities, and receiving treatment, while women in high CS clusters face minimal barriers and may access CS even when not strictly indicated.

Our finding of an 8.4-fold variation in CS rates across clusters exceeds the approximately 5-fold variation reported in wealth stratified analyses from previous Nigerian studies. This suggests that cluster analysis, by combining multiple dimensions simultaneously, reveals greater heterogeneity than single variable stratification. The universal presence of Cluster 9 across all geopolitical zones at 5.9% to 15.0% indicates that urban populations with moderate socioeconomic characteristics exist throughout Nigeria, likely concentrated in state capitals.

Several limitations warrant consideration. The substantial sample reduction from 39,050 to 13,915 women primarily reflects restriction to those with recent births and may introduce selection bias. The silhouette score of 0.227 indicates moderate cluster quality with some overlap between clusters, and the dataset lacked information on clinical indications for CS, limiting our ability to distinguish medically indicated from potentially elective procedures.

## 5.0 CONCLUSION

This study applied K-means cluster analysis to identify distinct obstetric risk profiles associated with caesarean section utilization among 13,915 Nigerian women with recent births. Ten clusters were identified with CS rates ranging from 1.7% to 14.4%, revealing an 8.4-fold disparity that underscores profound inequities in access to this life saving intervention.

The findings demonstrate clear geographic segmentation, with North Central and North East zones dominated by the lowest CS clusters at rates of 1.7% and 2.3% respectively, while South West and South South zones were dominated by the highest CS clusters at rates of 14.4% and 10.4% respectively. Over half the study population belonged to clusters with CS rates below 2.5%, indicating substantial unmet need for emergency obstetric care concentrated in rural Northern Nigeria.

Based on these findings, we recommend that policymakers prioritize emergency obstetric care investments in North Central and North East zones where over 60% of women belong to severely underserved clusters. Program implementers should develop geographically differentiated strategies, expanding facility capacity and emergency referral systems in rural areas while promoting evidence-based CS indications in urban Southern regions. Healthcare providers should strengthen antenatal care as a platform for identifying high risk pregnancies and ensuring timely referral.

Future research should conduct longitudinal studies linking cluster membership to maternal and neonatal outcomes, incorporate clinical indication data to distinguish appropriate from potentially inappropriate CS utilization, and validate the cluster solution across multiple survey rounds to assess temporal stability. Machine learning based clustering provides a powerful tool for unmasking hidden disparities, and the profound inequities revealed demand urgent attention to ensure all Nigerian women can access caesarean section when needed while avoiding unnecessary surgical intervention when not indicated.

### Ethics declaration

This study is a secondary analysis of publicly available, de-identified data from the Nigeria Demographic and Health Surveys (NDHS) for 2024. The original surveys were conducted by the National Population Commission (NPC) of Nigeria in collaboration with the DHS Program, following strict ethical protocols including informed consent and institutional review board approvals. As this research utilized anonymized secondary data, further ethical approval was not required for this analysis.

### Consent to participate declaration

Not applicable. This study used secondary, de-identified data from the Nigeria Demographic and Health Survey (NDHS); the original data collection process by the DHS Program included obtaining informed consent from all individual participants.

### Consent to publish declaration

Not applicable. The manuscript does not contain any individual person’s identifiable data, images, or videos.

## Clinical trial number

Not applicable.

## Data availability statement

The datasets analyzed during the current study are available from the DHS Program website (https://dhsprogram.com/) upon registration and approved request.

## Authors contributions

**Samuel Ayoola Ajeboriogbon** conceived and designed the study, developed the methodological framework, and led the implementation of the K-means clustering algorithm, including data preprocessing and the optimization of cluster selection through validation indices. **Abolaji Moses Ogunetimoju** performed the advanced statistical modeling and Principal Component Analysis, interpreted the clinical risk profiles, and conducted the analysis of distribution across geopolitical zones. **Oluwafemi Lawal Bisiriyu** provided critical oversight, conducted the final editing, and performed a comprehensive review of the manuscript. All authors contributed to the drafting and critical revision of the manuscript, synthesized the policy implications, and approved the final version for submission.

## Institutional Review Board Statement

This methodological study utilized de-identified, publicly available secondary datasets. As such, no additional ethical approval was required beyond the original data collection protocols.

## Data Availability

All data produced in the present study are available upon reasonable request to the authors

## Acknowledgment

The authors would like to express their gratitude to The DHS Program (ICF International, Rockville, MD, USA) for granting access to 2024 Nigeria Demographic and Health Survey (NDHS) datasets used in this study. We also acknowledge the National Population Commission (NPC) of Nigeria for their role in the successful implementation of the surveys and for ensuring the availability of high-quality, nationally representative data. Special thanks are extended to the fieldworkers and the thousands of women across Nigeria who participated in these surveys, providing the essential data that makes this reproductive health research possible. Finally, we thank the reviewers for their insightful comments that helped refine the K-means clustering analysis presented in this work.

